# Detailed evaluation of chromatic pupillometry and full-field stimulus testing to assess ultra-low vision in retinitis pigmentosa

**DOI:** 10.1101/2022.09.09.22279766

**Authors:** Midori Yamamoto, Take Matsuyama, Tadao Maeda, Seiji Takagi, Naohiro Motozawa, Daiki Sakai, Yasuhiko Hirami, Akiko Maeda, Yasuo Kurimoto, Masayo Takahashi, Michiko Mandai

## Abstract

**Purpose:** Novel therapeutic options, such as regenerative medicine and gene therapy are now emerging as viable treatment options for patients with severe visual impairments, such as retinitis pigmentosa (RP). Gradable assessment of patients’ visual function is essential to consider treatment options and to evaluate treatment outcomes, however, evaluation of visual function in advanced low vision patients is often challenging due to patients’ poor and sometimes unpredictable responses. In this study, we attempted to accurately assess the visual capabilities and disease stage in RP patients with visual acuity of 0.01 or lower, using chromatic pupillometry and full-field stimulus testing (FST), combined with retinal structural features, as determined by spectral-domain OCT.

**Design:** Retrospective analysis of visual function tests in 84 eyes of 43 patients with advanced RP with visual acuity of 0.01 and lower visited Kobe City Eye Hospital from 2019 to 2021.

**Methods:** Eighty-four eyes of 43 patients with advanced RP with visual acuity of 0.01 and lower were evaluated by chromatic pupillometry, FST, BCVA (best-corrected visual acuity), and OCT (optical coherence tomography). Hierarchical (multilevel) Bayesian modeling was used to estimate individual eye’s pupil response and FST, taking into account the ambiguity and randomness often observed in ultra-low vision patients. Using the estimated ability obtained from each test, the correlation between each test and retinal thickness was further analyzed to make a comprehensive assessment of the data.

**Results:** FST and pupillometry measurements were moderately correlated with visual acuity, but exhibited a wide range of values within the same visual acuity groups. FST was not correlated with central retinal thickness at CF/HM VA range and seemed to reflect overall remaining photoreceptor function including peripheral retina. Pupillometry seemed to distinguish different levels inner retinal function.

**Conclusion:** The combination of pupillometry and FST allowed for graded evaluation of visual function within patients grouped in the same visual acuity groups in advanced RP patients with ultra-low vision.

## Introduction

Retinitis pigmentosa (RP) is an inherited retinal degeneration with a reported prevalence of 1 in 4000 and is one of the leading causes of severe visual dysfunction worldwide.^1^ In typical retinitis pigmentosa, the preceding loss of rod photoreceptor cells leads to night blindness and concentric visual field constriction, followed by a progressive loss of cone photoreceptor cells resulting in deterioration of central vision, sometimes leading to blindness.

As with many neurodegenerative diseases, there have been no established treatments to restore visual function once it is lost. However, in recent years, the development of retinal cell-based therapies and gene therapies have been put in clinical trials for severe retinal diseases. These new therapies conceptually allow for high expectations that they will not only inhibit the disease progression but also improve visual function.

Optogenetic intervention, which aims to restore light responsiveness by ectopic expression of light activated proteins in inner retinal cells, is an alternative gene therapy approach which have also been applied to human patients^2^. Regenerative medicine is a replacement therapy for damaged or lost cells to regenerate visual function. We reported a proof-of-concept for pluripotent stem cell (PSC)-derived retinal organoid transplantation in animal models of RP by confirming host-graft synapse formation, restoration of RGC light responses after transplantation, and also by behavioral improvement^3,4^. Based on these POC, we recently conducted a clinical trial for RP patients using human-induced pluripotent stem cell (iPSC)- derived retinal organoids (jRCTa050200027)^5^.

Development and clinical application of these new therapies mandate an accurate and gradable assessment of patients’ visual function, since they target specific points of the visual pathway and disease stage. Unfortunately, patients with “severe visual dysfunction”, for example patients with a visual acuity lower than 0.01, which are the primary candidates for these new treatments, are often grouped together into broad categories. A more detailed assessment of which part of the visual pathway have a residual function, and to what extent, will inform the best treatment options. If the therapeutic concept is to enhance photoreceptor function through gene therapy or other means, it is important to determine the degree of photoreceptor survival in the retina. For photoreceptor transplantation, it is crucial that secondary and tertiary neurons that transmit signals from photoreceptors are functioning, or can become functional following photoreceptor transplantation. Optogenetics and artificial eye approaches would require a functioning neural pathway from retinal ganglion cells to the visual cortex. A more accurate assessment of visual function and disease stage will help determine the indications and methods of treatment. Additionally, the same methodology could be utilized to reliably determined the effectiveness of the treatment and assess the extent of improvement in visual functions, if any.

To assess visual function in late-stage RP patients with low vision, a number of functional tests have been reported including full-field stimulus testing (FST) ^6−8^, and chromatic pupillometry^9−12^. FST is a subjective test using the full field light stimuli, which can theoretically separate the cone and rod photoreceptor responses by using different wavelengths of the stimulus. Chromatic pupillometry is a noninvasive and objective method to study pupillary responses induced by cone, rod photoreceptors and melanopsin-containing intrinsically-photosensitive retinal ganglion cells (ipRGCs), which can be distinguished by the intensity and wavelength (color) of light stimuli. ^11,13,14^ Light information for pupillary light reflex is relayed to the brain through a pathway separate from the conventional visual pathway, with optic tract fibers terminating at the pretectal nucleus in the midbrain, and not at the LGN of the thalamus.^15−20^ Selected stimulus conditions can produce pupil responses that reflect phototransduction primarily mediated by rods, cones, or melanopsin/ipRGCs. We conducted a detailed analysis of FST and chromatic pupillometry on advanced RP patients with ultra-low vision. We then examined the correlation between the results of these tests and conventional visual acuity (VA) test, as well as OCT observation. Our results indicate that chromatic pupillometry and FST together with OCT can be used complementarily to more accurately evaluate the visual function in advanced RP patients and make better assessments on estimated pathological status of patients.

## Methods

### Patients

This retrospective observational study protocol was approved by the institutional review board of Kobe City Medical Center General Hospital (approval number: E18006). The study was conducted in accordance with the terms of the Declaration of Helsinki. Written informed consent was obtained from all participants after an explanation of the nature and possible consequences of the study. We included the patients with both RP and RP-related diseases who visited the Kobe City Eye Hospital, Kobe, Japan between 2019 and 2021. Inclusion criteria for patients were that the visual acuity was equal to or lower than 20/2000. Examinations were performed in 84 eyes of 43 subjects (Table 1) and all patients underwent a series of ophthalmic examinations that included the following: best-corrected visual acuity (BCVA), intraocular pressure (IOP), fundus examination, spectral-domain optical coherence tomography (SD-OCT), chromatic pupillometry and FST.

**Table 1.**
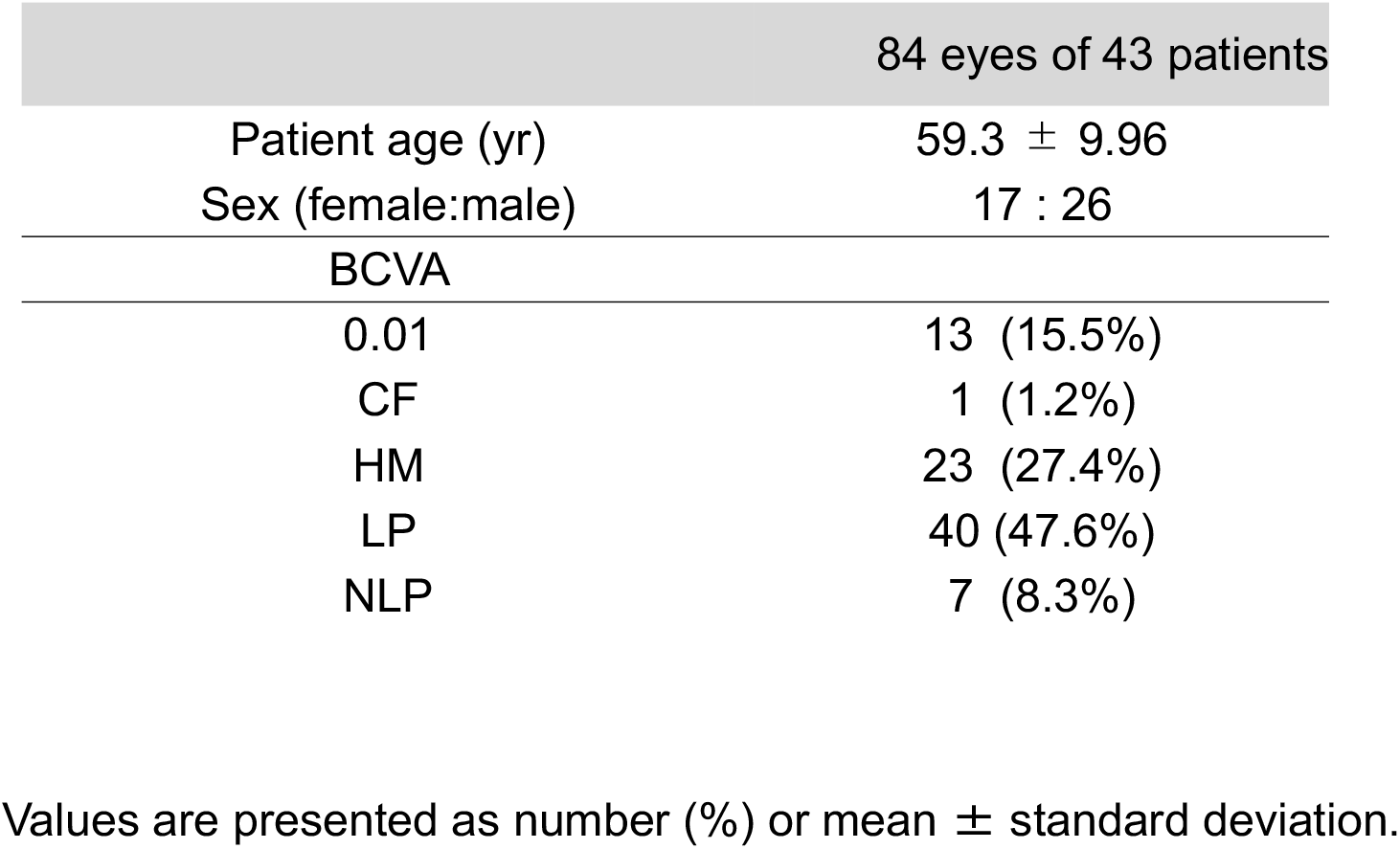
Clinical characteristic of the patients

### Visual acuity measurements

BCVA was tested using a standard Landolt C acuity chart. The visual acuities were converted to logMAR to allow for statistical analysis. Visual acuity of 0.01 were assigned logMAR visual acuity of 2.0, counting fingers (CF) 2.6, hand motion (HM) 2.9, light perception (LP) 3.1, and no light perception (NLP) 3.4, following the convention of Jhonson et al.^21^

### SD-OCT evaluation

The SD-OCT images were obtained by using the Heidelberg Spectralis (Heidelberg Engineering, Spectralis, Heidelberg, Germany). The thickness of total retinal thickness(TRT) and the thickness of Ganglion Cell Complex (GCC), Inner Nuclear Layer (INL), and photoreceptor layer (photo) were measured at four cardinal points 2,000 µm from the fovea in horizontal and vertical scans. All measurements were performed by using with “caliper” function of the Heidelberg instrument. TRT was defined as the distance between the signal peak at the vitreoretinal interface (the internal limiting membrane: ILM) and the posterior boundary of the major signal peak that corresponds to the basal retinal pigment epithelium/Bruch’s membrane complex (RPE/BrM). GCC was defined as the three innermost retinal layers: the nerve fiber layer, the ganglion cell layer, and the inner plexiform layer. INL was defined as the distance between the basal Inner Plexiform Layer (IPL) and the OPL. Lastly, the photoreceptor layer was defined as the region between Outer Plexiform Layer (OPL)and RPE/BrM.

### Chromatic pupillometry

Chromatic pupillometry and FST measurements were performed using an Espion system with a ColorDome light stimulator in a completely dark room. The protocol for chromatic pupillometry consisted of the following four steps. For each step measurements were repeated 5 times. The first step (rod) consisted of a -3 log cd/m^2^ blue light stimulus following 10 minutes of dark adaptation. The second step (cone1) consisted of a 1 log cd/m^2^ red light stimulus on 0.1cd/m^2^ blue background light following 2 minutes of light adaptation. The third step (cone2) consisted of a 3 cd/m^2^ white stimulus light on 1 cd/m^2^ white background light following 2 minutes of light adaptation. Lastly, the fourth step (mela) consisted of a blue light stimulus of 150 cd/m^2^ without background light following 2 minutes of dark adaptation. Rod and cone responses are observed as transient responses immediately after light stimulation whereas melanopsin response is observed as a sustained pupil response. Each test was performed on both eyes simultaneously, and changes in pupil diameter were recorded with an infrared camera in the color dome™.

### Full-field stimulus testing (FST)

FST was examined using the same equipment as chromatic pupillometry. After 45 minutes of dark adaptation following mydriatic eye drops in both eyes, blue (448nm), white (590nm), green (530nm), and red (627nm) light stimuli were used to examine the rod and cone functions. The non-tested eye was shielded so that one eye was tested at a time. Subjects were given a button box (Diagnosys LLC, MA, USA) with a Yes/No button and answered whether they perceived light in the color dome™ after a beep. Thresholds to the light stimulus were measured for each color while also taking into account false negatives and false positives.

### Statical analyses

We used full Bayesian statistical inference with MCMC sampling for statistical modeling using Rstan (Stan Development Team. 2017. RStan: the R interface to Stan. R package version 2.21.5. http://mc-stan.org). Since most data consists of measurements gathered from both eyes of subjects, we incorporated terms for the patient and eye bias to account for the natural hierarchical structure of the data. Using these and other relevant parameters, such as color of stimulus light, we obtained posterior estimates from linear regression models. We show the posterior distributions of parameters of interest, which indicate the probability for the value of the parameter given the data, with 95% compatibility intervals (confidence intervals) and mean indicated. Note that posterior distributions do not represent a simple pooling (average) of data for a particular set of predictor combinations, rather they represent the effect of predictors while considering the data as a whole. Detailed explanation of the statistical models, as well as the Stan code for the models are provided in a GitHub repository (https://github.com/matsutakehoyo/Clinical-Data-Analysis).

## Results

### Robust hierarchical modeling of FST makes better threshold estimates

FST measurements were analyzed using a multilevel (hierarchical) logistic model as illustrated in Figure 1A. We estimated patients’ ability to detect light taking into consideration the effect of light intensity, (*x*_*i*_, in dB), patient (*β*_*pat*_), eye (*β*_*eye*_), and stimulus color (*β*_*clr*_) as predictors in our model. We also allowed eye biases to vary for different colors (eye and color interaction, denoted eyeXclr). Figure 1B shows the posterior distribution of model parameters. Overall, despite the presence of a large patient to patient variation (*β*_*pat*_), most of the patients showed little variation between the responses by right (RE) and left eyes (LE). There is a clear effect for the color of the stimulus light, with patients being most sensitive to blue light than red light (Figure 1B). A key aspect of the model is the inclusion of the ‘guessing’ parameter (*α*) for robustness. The ‘guessing’ parameter is a measure of points that do not conform well with the logistic regression, i.e. the fraction of outliers. The small ‘guessing’ parameter estimates (mode < 0.05) for most eyes support the reliability of this test, with some notable exceptions where subjects seem to be ‘guessing’ more than half of the time (Figure 1B). Figure 1C shows representative cases with posterior predictions from our model alongside data measurements with the ‘guessing’ rate presented on top of each panel. Typically, a subject’s probability of correctly identifying the light stimulus decreases as the light intensity (*x*_*i*_) decreases, as described by the S-shaped curve of the logistic function. Vertical lines show the threshold value (the light intensity at which the probability of success equals 0.5). The sensitivity to color generally decreases from Blue to Red, although the amount of the shift varies from patient to patient and even from RE to LE (id =4 and id =28 for example). In samples with larger ‘guessing’, subject’s responses were seemingly random and measurements tended to fall outside the S-shaped curve of the logistic regression (patient id = 16 and 28 RE). Although the difference in threshold values between our estimates (solid line) and those made by the measurement instrument (dotted line) are practically identical in most of the cases, there are also cases where they differ significantly (id= 16). The dotted line is estimated from the measurements for a particular sample (eye) at a particular color in isolation (no pooling), whereas our estimates take into account all of the data, from all conditions and from all samples (hierarchical partial pooling). In hierarchical partial pooling, estimates are more skeptical of measurements that fall outside the group means. For example, the instrument estimated threshold (dotted line, no pooling) would indicate that id = 16/RE has much higher threshold for Blue light than for Red light, which is physiologically unlikely as the response to Blue stimuli includes contributions from both rods and cones, whereas responses to Red stimuli is solely driven by red cones. The hierarchical partial pooling estimate is instead largely informed by the overall color effect (*β*_*clr*_), which places Blue at lower thresholds than Red, and patient and eye biases (*β*_*pat*_, *β*_*eye*_). The large ‘guessing’ is also an important factor here, as some of the observations are seemingly random. As these estimates are not strongly supported by the data, compatibility intervals encompass a wider range of values. These estimates are nonetheless useful as they narrow the reasonable and credible range of values. Similarly, using the available information, we are able to make inferences even if data is entirely missing for a particular color (id = 3 Blue, id = 30 Green for example), or in cases where no correct responses were obtained within the tested range (id = 6 LE). Overall, we believe our model is able to capture and evaluate the uncertainty in measurements, giving us estimates with reliable confidence intervals reflective of the subject’s true ability to detect light.

**Fig. 1.**
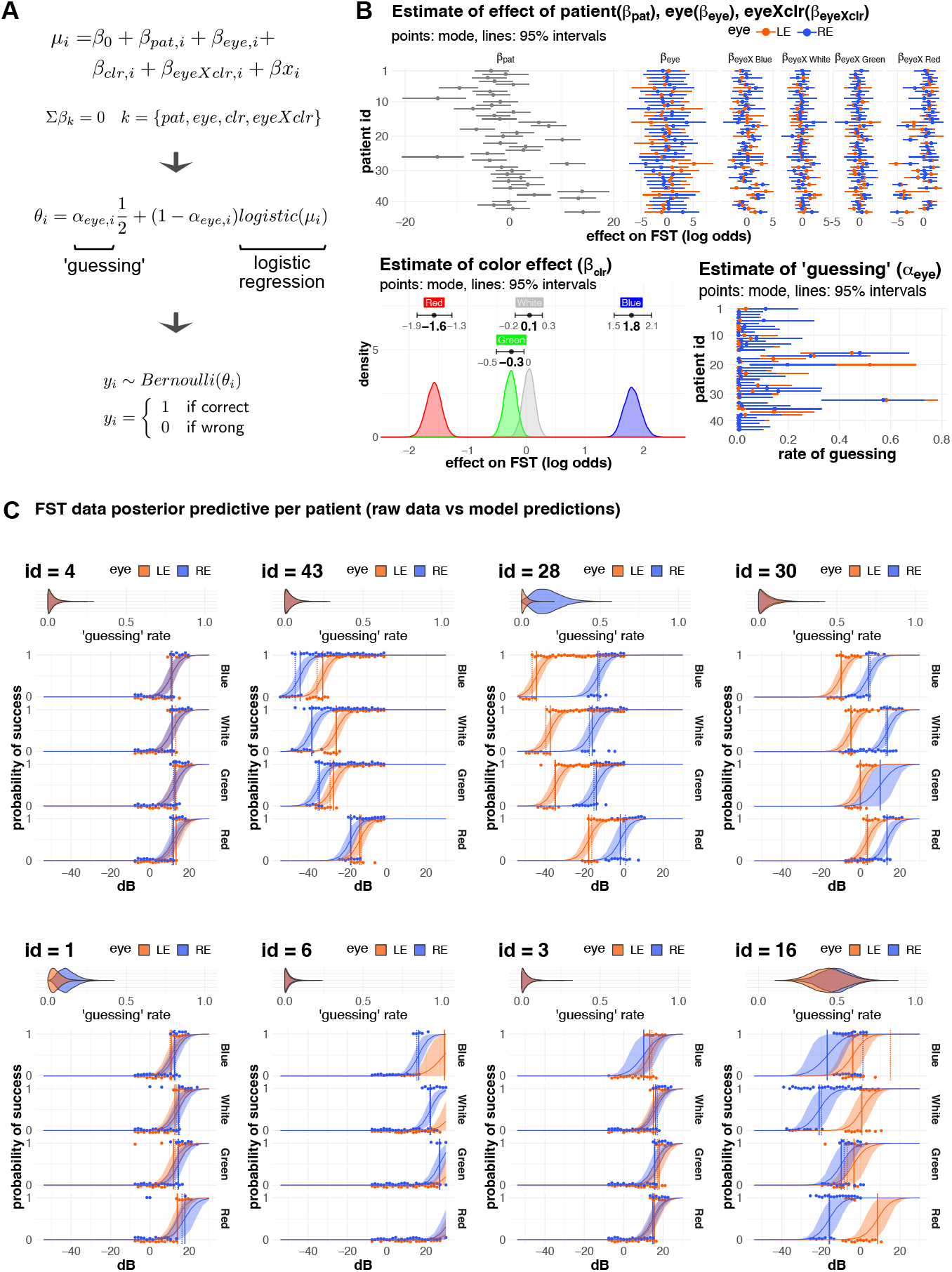
Analysis of FST A. FST Model. Patient’s responses *y*_*i*_ (correct or wrong) to a light stimulus were analyzed with a logistic regression model. In a typical FST experiment, the intensity of the light stimulus (*x*_*i*_, measured in dB) is gradually decreased as the subject pushes a bottom if they can perceive the light. We used multilevel (hierarchical) modeling, to estimate the effect of different light stimulus color (*β*_*clr*_), patient (*β*_*pat*_), and patient eye (*β*_*eye*_) on the probability of correctly detecting the light stimulus. In order to allow higher freedom for estimates of each eye to vary at different light color we added an interaction term for patient eye and light color (*β*_*eyeXclr*_). Each parameter was constrained so that Σ*β*_*k*_ = 0 (sum to zero constrain) and the effect of each parameter is shown as deviation from the overall mean (*β*_0_). The probability of seeing the light stimulus is given by the logistic model and a ‘guessing’ parameter (*α*), which is estimated for each eye. This parameter accommodates data points that do not conform well with the logistic regression. B. Posterior estimates of FST model. Estimates for patient (*β*_*pat*_) vary widely whereas *β*_*eye*_ and *β*_*eyeXclr*_ variation is relatively smaller. Most of the patients have similar estimates for LE and RE although some patients seem to have a significant LE/RE difference. There was a clear effect for the light color, with subjects being more sensitive to blue light than red light (Red < Green, White < Blue). Although most of the subjects had a relatively small ‘guessing’ estimate (*α*) with the mode and 95% interval below 0.1 (=10% guessing), there were notable exceptions with very large guessing values (mode > 0.2). C. Examples of raw data and model estimates. Points show the raw data for a series of experiments for a patient. Note that points are shown slightly offset from their true values (0 or 1) to avoid RE and LE points overlapping. Model estimates are shown with a solid curved line (mean) and ribbon (95% intervals). ‘Guessing’ estimates are shown as violin plots on top of the FST results. The vertical lines show the FST threshold values (dB at which probability of success is 0.5). The solid line represents the mean threshold value of our estimates, whereas the dotted line shows the threshold estimated by the measurement apparatus (threshold estimated individually). Color indicates LE (orange) and RE (blue).

### Analysis and characterization of pupil responses in eyes with advanced RP

Pupillometry measurements were analyzed using a multilevel (hierarchical) multivariate model as illustrated in Figure 2A. Measurements consists of time series of the pupil diameter changes to different stimulus (stimulus is applied at t=200 ms). For each stimulations condition (rod, cone1, cone2, and mela) there are 5 repeats. While repeated measurements seem to reliably reproduce pupil diameter changes for rod, cone1, and cone2 stimulations, this was not the case for mela stimulation. In most cases, the first repeat elicited a large reduction in pupil diameter, while repeats 2-4 exhibited a more attenuated response. We therefore separated the response to mela stimulation to mela1 and mela2 in order to differentiate between the first and the remaining repeats. Although the data is a time series, we focused on three key regions: before, peak, and after light stimulation, to simplify the analysis. We extracted the pupil diameters from these regions for each measurement and estimated their values. Figure 2B shows the posterior distribution of some key parameters. We used a multivariate distribution as before, peak, and after pupil diameter values tended to be highly correlated. We allow every eye and light stimulation condition to vary while estimating the bias for each subject and the effect of light stimulation. Most of the variance in the data is explained by patient-to-patient variation (*β*_*pat*_ > *β*_*eye*_, *β*_*eyeXclr*_), although substantial LE/RE differences were observed in some cases. Figure 2C shows the expected typical response (*β*_0_ + *β*_*clr*_). Note that peak is not defined for mela1 and mela2, as we do not expect, or observe, a sharp peak for these. Rod, cone1, cone2, and mela1 measurements have a baseline (before) of about 5.5 (Rod: 5.6mm, Cone1/Cone2: 5.2) mm. There is very little or no pupil change to Rod stimulation as is expected in RP patients.

**Fig. 2.**
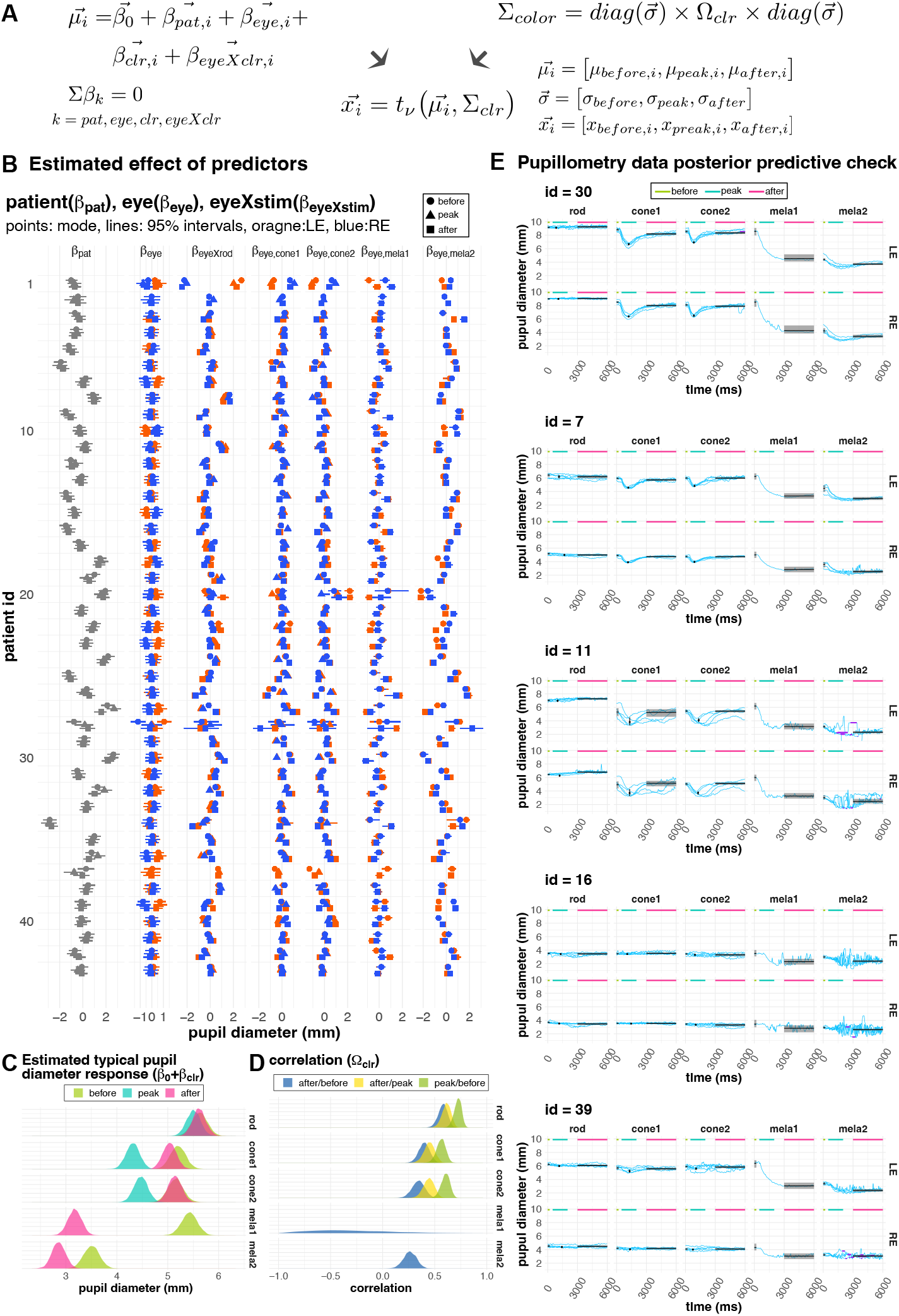
Analysis of chromatic pupillometry A. Pupillometry model. Before, peak, and after were defined as the pupil diameter values before light stimulation (0 − 200 ms), peak value following light stimulation (200-2000 ms, defined only for rod, cone1, and cone2 stimuli), and value of the relatively flat region following light stimulation (3000 − 6000 ms) respectively. These pupil diameter measurements (*x*_*before*_, *x*_*peak*_, *x*_*after*_) were modeled as a multivariate distribution, taking into account their inter-correlation. We used the *t*-distribution for robustness since some values may have been influenced by measurement artifacts (see Methods for details). Note that de degrees of freedom *v* is also a parameter that is estimated from the data. Similar to the FST analysis, we incorporated patient (*β*_*pat*_), eye (*β*_*eye*_), color (*β*_*clr*_), and an interaction term for eye and color (*β*_*eyeXclr*_) to allow estimation of overall trends while allowing for flexibility to account specific trends for each eye/color combination if there is strong evidence. Parameters were constrained so that Σ*β*_*k*_ = 0 (sum to zero constrain) and the effect of each parameter is shown as deviation from the overall mean (*β*_0_). The correlation between measurements was estimated for each stimulation condition (*ρ*_*clr*_). B. Posterior estimates of PM model. Estimates for effect off *patient, eye, eyeXclr* are shown per patient (rows). Note that for each patient id there are three estimates for before, peak, and after (shown as different shapes). C. Posterior distribution of predicted typical response (*β*_0_ + *β*_*clr*_). D. Posterior estimates of correlation coefficient for each light stimulus. E. Examples of raw data and model estimates. Light blue traces show the individual pupillometry measurements. Purple dots overlayed on the blue traces (id =16 and 39) show data points that were excluded from the analysis, as they consist of regions with constant values (sd = 0) and therefor likely represent regions where pupil detection failed. Rod, cone1, and cone2 measurements consist of 5 traces (repeats), whereas the 5 traces of mela light stimulus were separated to mela1 (repeat 1) and mela2 (repeats 2-5), as we noticed that there was a large difference between mela1 and mela2. The black and gray shaded area show the estimated mean and 95% confidence interval for before, peak, and after pupil diameter.

Rod/Cone (cone1, cone2) stimulation induce a small change, with pupils transiently constricting to about 4.3-4.5 mm, and immediately reverting back to baseline values. On the other hand, there is a large constriction of the pupil for mela1 (3.2 mm) which only partially reverts (3.5 mm), resulting in very small pupil diameter change for mela2. Figure 2D shows the estimated correlation coefficients for the different light stimulation conditions (rod, cone1, cone2, mela1, and mela2). Before, peak, and after values had a high correlation for rod, cone1, cone2, and mela2 stimulations. This trend is disrupted for the correlation between before and after measurements for mela1, which show most of the posterior distribution with negative correlation values. This indicates that, pupil diameter changes in mela1 are larger when pupil diameter is larger before the light stimulus.

Figure 2E shows some of the data highlighting typical and noteworthy cases. Model predictions (posterior predictions = *β*_0_ + *β*_*clr*_ + *β*_*pat*_ + *β*_*eye*_ + *β*_*eyeXclr*_) reasonably describe the data. Figure 2E id = 30 shows a typical response, with an absent rod response, a small peak response for cone1 and cone2 stimuli, a large mela1 response (repeat #1) and a very small mela2 response (repeat#2-5). In most cases the repeats of mela2 were largely identical, however there were a few cases where melanopsin response seem to gradually decrease. For example, figure 2E id =7 shows an example where baseline values for mela2 gradually decreased (repeat 1 (mela1) > repeat 2,3 > repeat 4,5). Figure 2E id = 11 shows an example where cone response varies greatly. Also there appears to be a rapid oscillation of the pupil following mela2 stimulation. Figure 2E id = 16 shows an example where all responses were absent except for mela1. Figure 2E id = 39 is similar to id = 16 however there is a much more robust response for mela1, and there is a large difference between LE/RE.

### FST and Pupil response is largely uncorrelated to VA and retinal thickness

Having analyzed FST and pupil response separately, we next focused our attention on how these correlate to each other. Figure 3 shows the pair plot for some representative features. We select sex, age, VA(logMAR), total retinal thickness within macular area (2mm from central fovea), FST (White), pupil diameter before stimulation, and pupil change to cone1, mela1, and mela2 stimulation for this analysis. We used the threshold for white light stimulation for FST measurements as representative data for FST measurements, as correlation between FST threshold for different stimulus color (Red, Green, White, Blue) was extremely high (*ρ* > 0.9, data not shown) and using any color would have resulted in practically identical results. Note the wide distribution of values for total retinal thickness, FST, and pupil diameters within the same VA group (Figure 3 blue rectangle), indicating that individuals within the same VA group may exhibit a wide range of visual functions and retinal thickness. The correlation analysis shows that VA is not significantly correlated to retinal thickness, or pupil responses, but moderately correlated to FST ((Figure 3 orange rectangle, *ρ*_*FST/VA*_ = 0.42). Since central macular cone area is generally considered last to degenerate in eyes with RP, we initially expected that macular retinal thickness to reasonably correlate with measures of visual function (FST, or Pupil response). However, we only found a moderate correlation with pupil response (*Pupil*_*clone*1_) in these eyes with low vision. Retinal thickness also seemed to be correlated to subject’s sex, with female subjects exhibiting a slightly thicker retina.

**Fig. 3.**
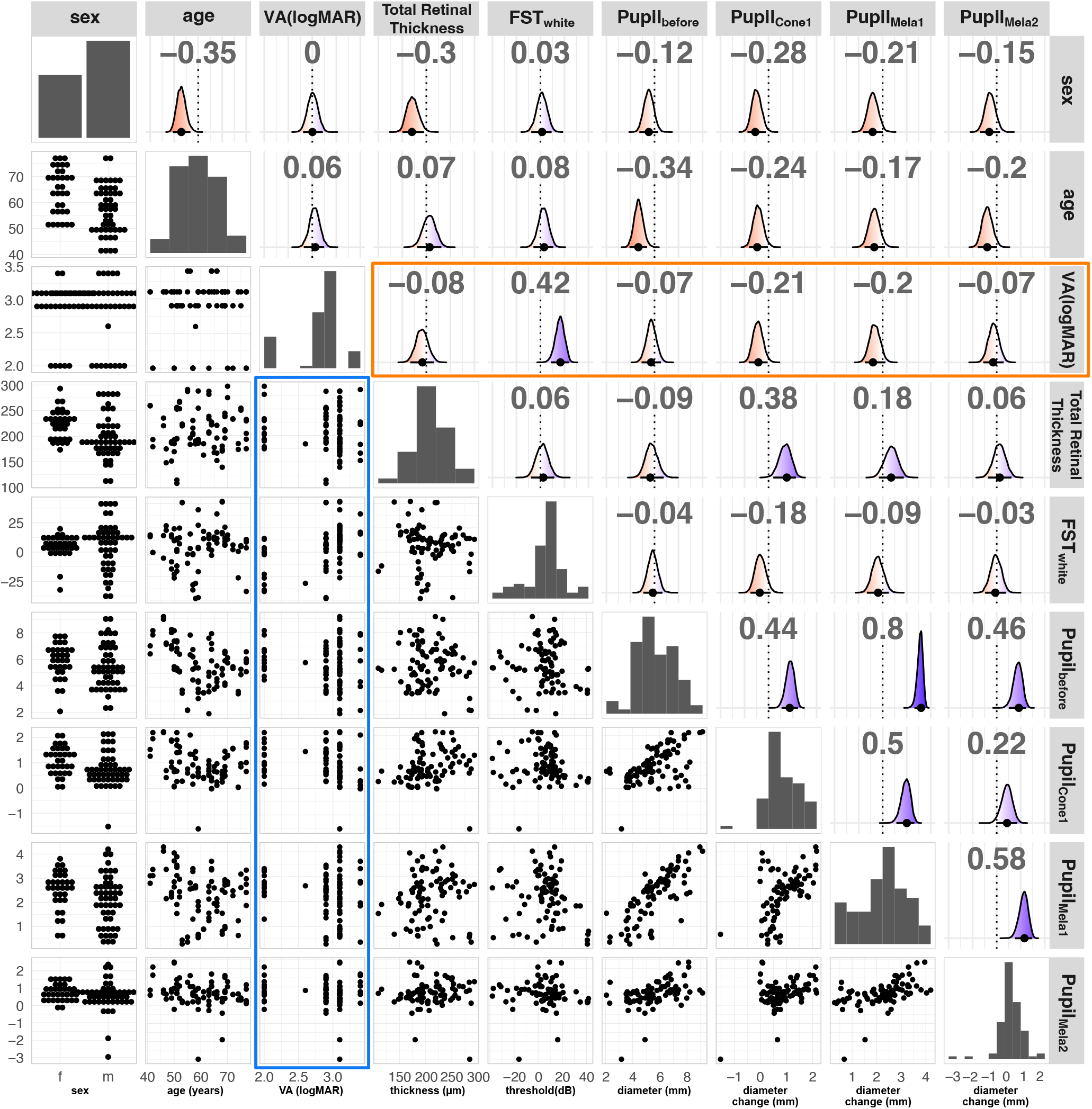
Pairwise relationship of dataset. We plotted (from left to right and top to bottom) sex, age, VA(logMAR), total retinal thickness (µ*m*), FST (db, white light stimulation), pupil diameter before light stimulation (*Pupil*_*before*_, mm), pupil change to cone1 stimuli (*Pupil*_*clone*1_, mm), pupil change to mela1 stimuli (*Pupil*_*mela*1_,*mm*), pupil change to mela2 stimuli (*Pupil*_*mela*2_, mm). The diagonal show distribution of each variable (bar chart for categorical data and histogram for continuous data). Lower triangular panels show the pairwise relationship of features. The blue rectangle highlights the relationship of VA(logMAR) to other measurements. Upper triangular panels show the estimated correlation coefficient, estimated from a multivariate model, with density plots showing the distribution of estimated correlation coefficients. Red indicates negative correlation (*ρ* = −1) and blue indicates positive correlation (*ρ* = 1). The mean (black point) and 95% confidence interval (black bar) are indicated on the bottom of the density plot. Numbers on top indicate the mean, and the dotted line denotes the zero line. The orange rectangle highlights the correlation coefficients of features with VA.

Figure 4 and 5 are per patient/eye summaries of collected data highlighting some characteristic cases. As we noted above, FST and pupil response does not correlate with VA or retinal thickness. The eye in Figure 4 shows that the retinal thickness and structure of the layer are relatively well preserved, with clearly defined ONL in fovea with visual acuity of 0.01. However, FST was poor and with a rather high guessing rate. We further looked into the Goldman visual field (VF) test to find that the eye had a very constricted central vision. In contrast, Figure 5 shows the eye with very thin retina in the macular area with visual acuity of HM. The eye however exhibited a relatively good FST with a clear shift between Blue and Red lights indicating rod functionality. Pupil response to rod stimuli was weakly present, supporting the presence of rod function in addition to cone function, which was rare in our RP patients. Goldman VF test show the presence of peripheral vision. Together these results indicate that both cone and rod function were preserved in spite of the very thin central retina.

**Fig. 4.**
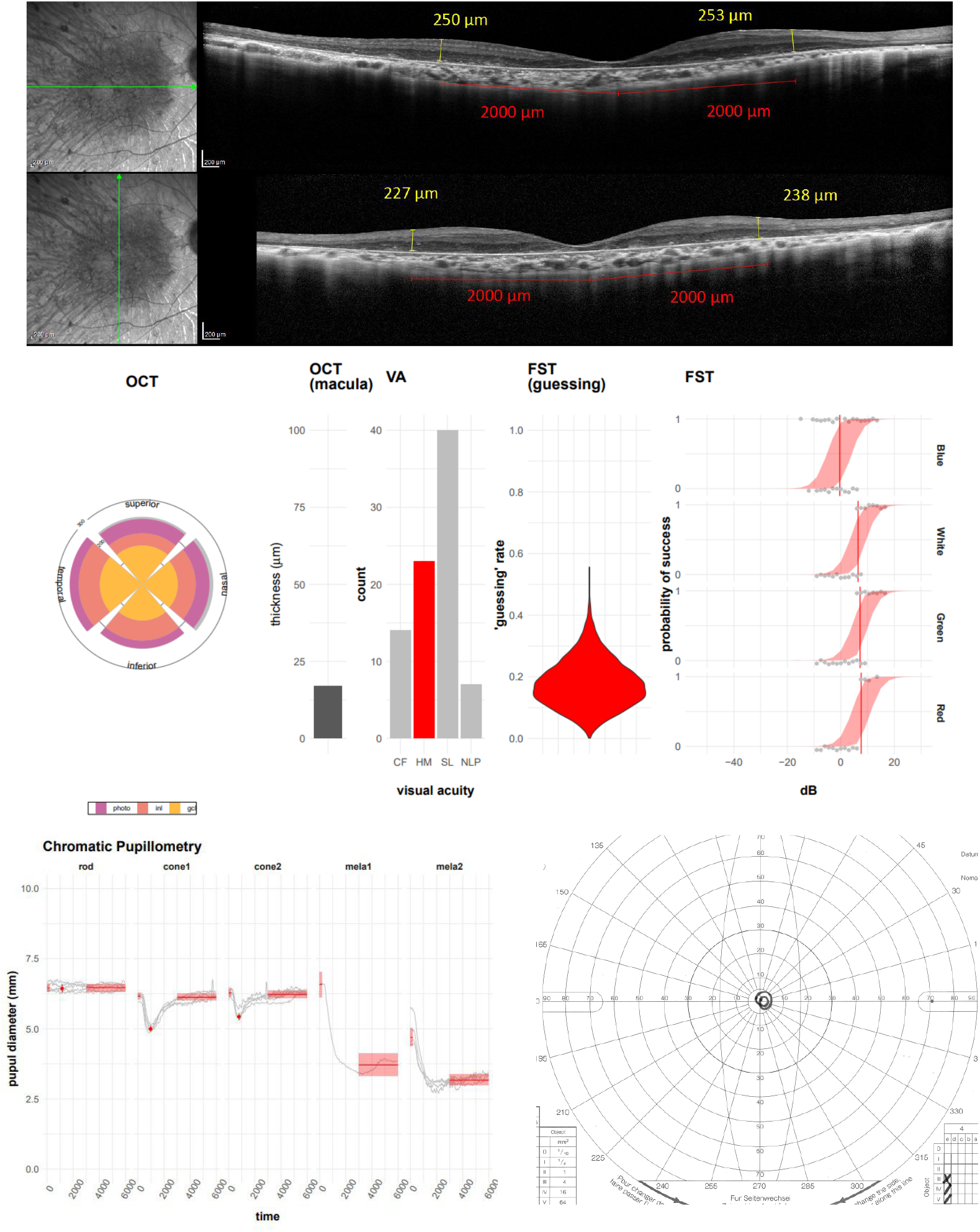
Representative case of an eye with a thick retina and poor FST. Vertical and horizontal OCT scans show ONL of the fovea is preserved and the structure of retina is appears to be well preserved. The average retinal thickness is near the macula was 242µm, which is equivalent to that of normal subject. The eye had been diagnosed as HM (center of middle panel). All of the pupillay responses (right of lower panel) except for rod response were clearly present. On the other hand, FST (right of middle panel) was relatively poor with threshold values over 0dB. The result of Goldman VF test (left of lower panel) shows a very constricted VF remained in the center.

**Fig. 5.**
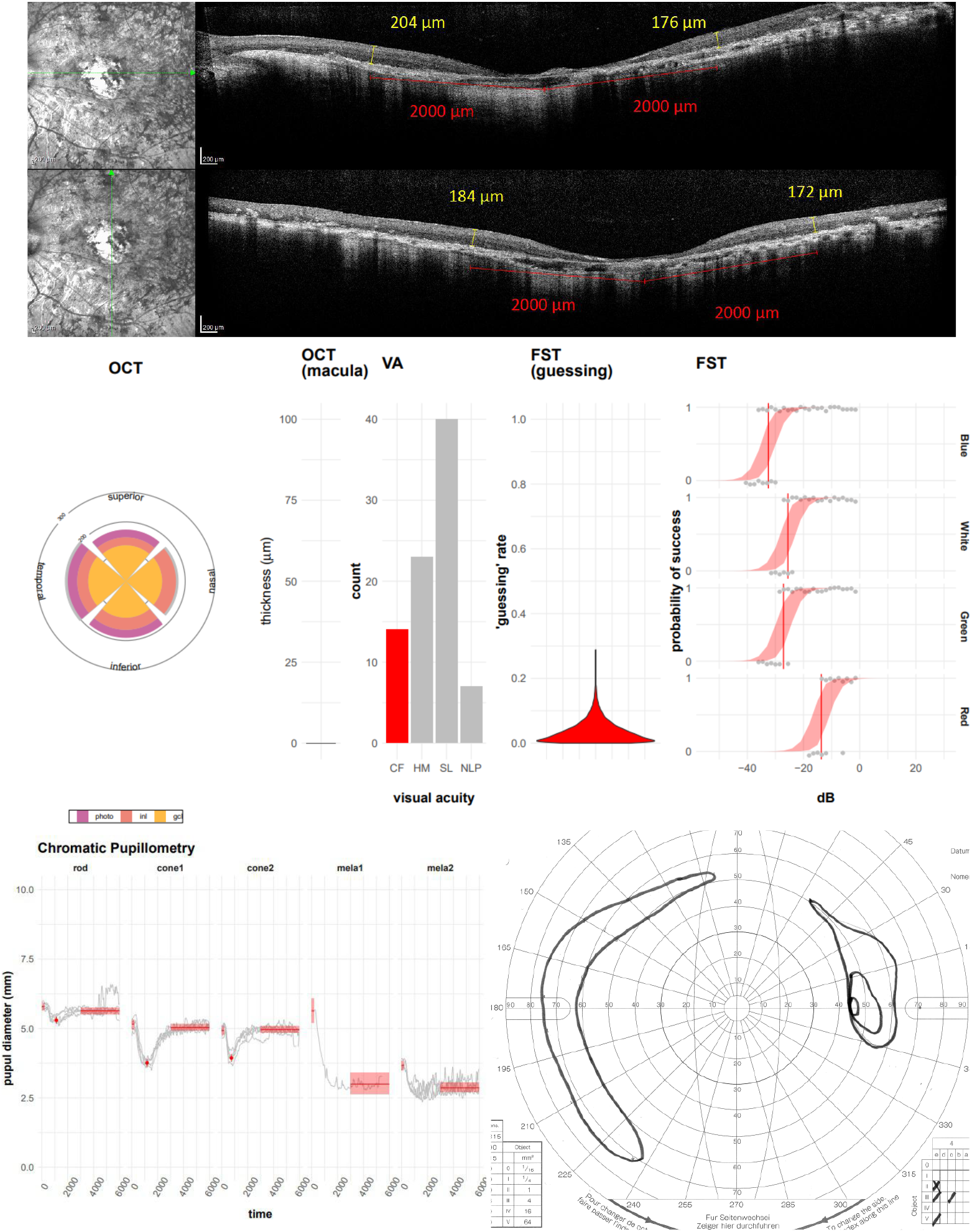
Representative case with thin retina but good visual acuity and FST sensitivity. The average of retinal thickness was 184µm and OCT images (upper panel) reveal photoreceptor cell loss and lack of fovea. VA (center of middle panel) was CF, the highest category of this study, and the FST sensitivities were similar to that of normal eyes. The response to chromatic pupillometry (left of lower panel) were also good (even rod response were confirmed which was rare in our study cohort) and GoldmanVF test (right of lower panel) shows a relatively large residual field of view in the periphery.

## Discussion

Birch et al. performed an FST study in inherited retinal disease(IRD)patients with visual acuity of 0.25 or better and reported that in some cases the threshold difference between blue and red stimuli was >10 dB.^22^ The decrease in the difference between Blue and Red thresholds is generally thought to reflect the more advanced retinal degeneration status.^6−8^Although our study cohort consisted of patients with more advanced retinal degeneration than that of Birch et al., we were nonetheless able to detect differences between Blue and Red FST (18 eyes (21%) (0.01/CF: 6 eyes, HM: 6 eyes, LP): 5 eyes, NLP: 1 eye).

Furthermore, we noted that eyes that have low threshold values tended to have a larger shift between Blue and Red light than eyes that have high threshold values (compare for example Figure1C id=43 and 3, note that higher threshold values indicate lower probability to respond to the light stimulus). To further understand the relationship between the shift in the threshold from Blue light, we plotted Green, White, and Red threshold values against Blue threshold values (Figure 7A). While Green and White values largely mirror Blue threshold values in all regions, red threshold values show a larger deviation from Blue values at lower thresholds. This is consistent with the notion that reduced sensitivity to Blue light represent advanced rod degeneration. Thus, while FST measurements for White or Green colors are highly correlated, Blue and Red FST values provide important diagnostic information in late-stage RP. Moreover, our data indicates that threshold values for White, Green, and Red light can reasonably be predicted from the Blue light stimulus alone, or vice versa.

OCT measurements, such as retinal thickness are widely and commonly used to diagnose retinal health, with well-organized thicker retinas generally presumed to correlate with more visual function. Since central cones are often preserved in RP, we measured retinal thickness within the macular area. However, we found no correlation between retinal thickness and indicators of visual function such as VA (logMAR), FST, or pupil response in our cohort of advanced retinal degeneration with visual acuity of 0.01 or lower (Figure 6). In order to further explore the relationship between OCT measurements and visual function we stratified the data into four groups according to subject’s visual acuity (0.01/CF, HM, LP, and NLP). Unexpectedly, we found a reversal of correlation between retinal thickness and FST values (Figure 6). Patients with thicker retina tended to have higher FST values (lower sensitivity for light perception) in 0.01/FC and HM whereas this trend is reversed in LP and NLP. Moreover, while Figure 6 shows the analysis using the average of 4 points for the entire retinal thickness (2,000 μm from the fovea), the same trend was observed at each of the quadrant position individually (temporal, nasal, superior, inferior) and for different layers (photoreceptor layer, INL, GCL), indicating that this is a global effect possibly affecting the entire macula. Although human visual function largely depends on central vision, example cases shown in Figure 4 and 5 suggest that in the eyes with advanced degeneration, remaining central visual function may be complemented by peripheral vision, possibly explaining the apparent discrepancy between the retinal thickness around the macula and overall visual function. It is of note that the FST and pupillometry measurements seemed sensitive enough to suggest the presence of remaining rod and cone function in the peripheral vision in the eye with “HM” (Figure 5). These results highlight that, together, FST, pupillometry and OCT measurements may help understand the pathological status of retinal degeneration, allowing for a more graded and comprehensive assessment than the conventional VA categories of FC, HM, LP, and NLP in ultra-low vision patients.

**Fig. 6.**
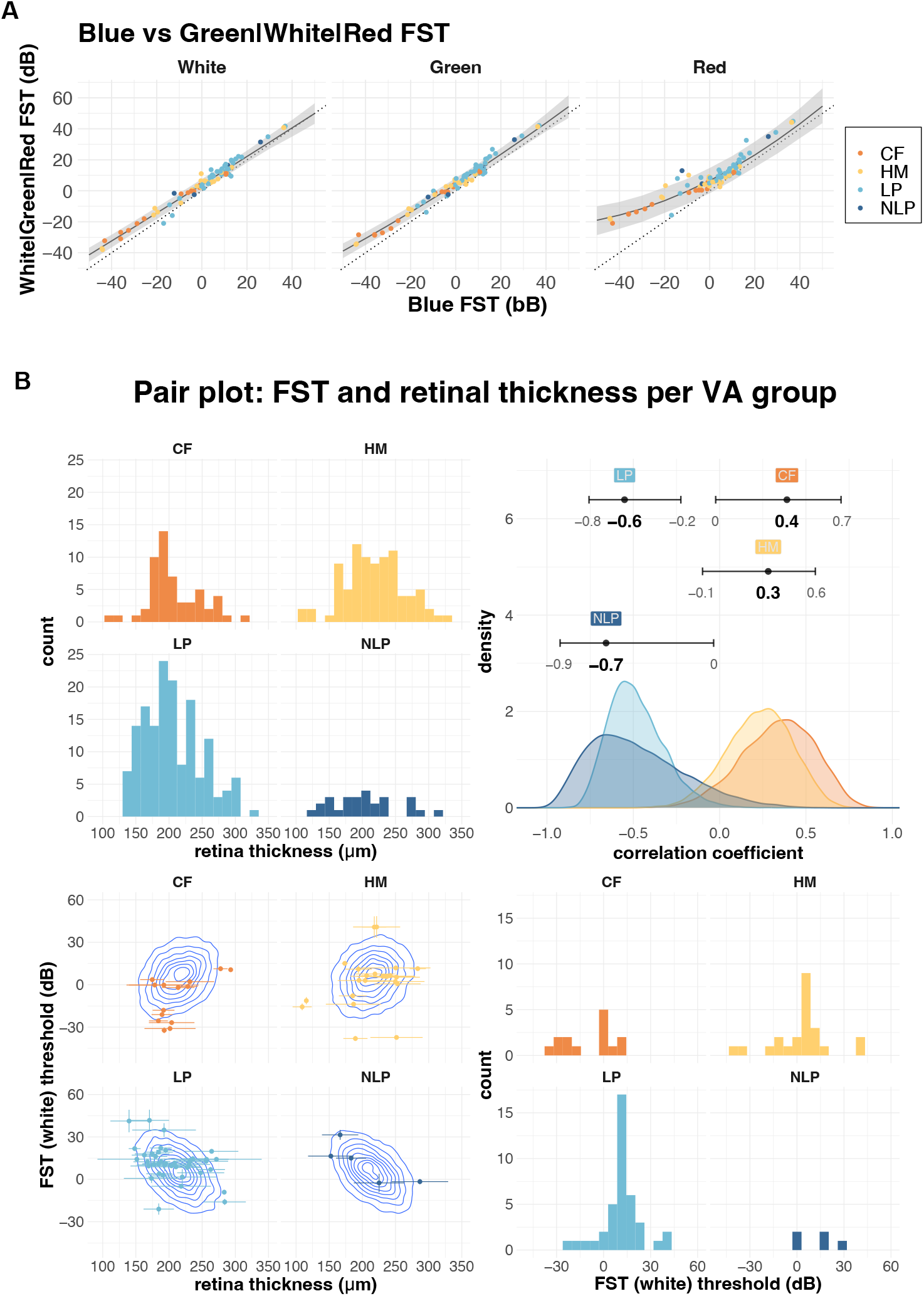
A. Relationship between FST values for Blue, White, Green and Red light. The Blue FST value was plotted against White, Green, and Red FST values. The dotted diagonal line has intercept = 0 and slope = 1. Points above the dotted line indicate higher FST value than Blue light and points below the dotted line indicate lower FST. The solid line shows the mean and the shaded area shows the 95% compatibility interval of quadratic regression for each of the colors. B. Relationship between retinal thickness and FST (white), stratified by four visual acuity groups. The upper left panel shows the distribution of retinal thickness and the lower right panel shows the distribution of FST for white light. The lower left panel show the distribution of the data (points) overlayed with estimated correlation (density plot), with horizontal error bars showing the SD of retinal thickness measurement (average of four points), and vertical error bars showing the estimated error (SD) from the FST analysis. The upper right panel shows the distribution of posterior estimates for the correlation between FST and retinal thickness (error bars show 95% confidence interval).

With the possibility of clinical application of novel treatments for visual restoration on the horizon, such as gene therapy, cell-based treatments, optogenetics, and prosthetic devices, a more detailed assessment of patient’s visual potency and the state of the retina is urgently needed. FST is very sensitive and has been reportedly used to detect subtle changes before and after treatment in a clinical study of gene therapy for LCA patients^8,23,24^. Indeed, in the current study, we obtained positive responses (defined as threshold below +15.0dB) from almost half of the eyes with “no light perception” highlighting the sensitivity of this approach. In the pupillary test, the two cone stimuli resulted in practically identical results and seem to reflect the remaining photoreceptor pathway and RGC activities. Positive FST and cone pupillary tests indicate a functional retinal circuitry even in the eyes with poor central vision. In these eyes, gene therapy or cell-based approaches may help enhance visual responses. On the other hand, eyes that only have a pupillary response to melanopsin stimuli, may better benefit from direct stimulation of RGCs such as artificial eye or optogenetic treatment rather than cell therapy. Lastly, eyes that completely lack FST and pupil responses may benefit from cortical interventions (Figure 7).

**Fig. 7.**
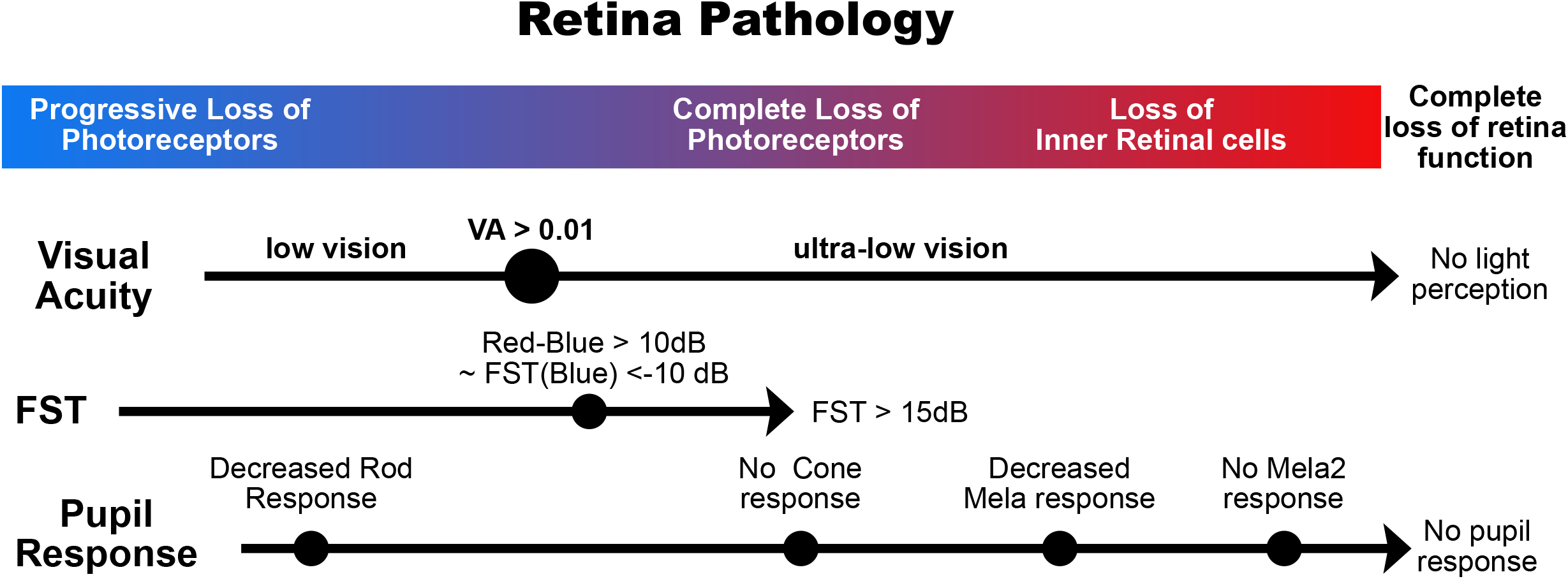
FST and pupillometry can be used, complementary to VA and OCT observations, to more precisely estimate pathological stage. As photoreceptors degenerate FST threshold increases. We use Red - Blue FST > 10bB as a reference point, indicating advanced rod loss. According to our analysis in figure 6A, this is equivalent to a blue FST < -10dB. The loss of photoreceptor cells in central macula as well as peripheral area leads to >15dB FST test results and the loss of pupillary cone response. Further degeneration and loss of retinal inner cells results in the decrease in pupillary melanopsin responses.

These emerging therapies for visual restoration bring hope for patients with severe visual impairments. The current study highlights the importance of developing diagnostic tools for ultra-low vision patients, bringing these treatments one step closer to clinical application.

## Data Availability

All data produced in the present study are available upon reasonable request to the authors

## Acknowledgments

We thank Dr. Yozo Miyake and Dr. Kaoru Fujinami for invaluable advice and all members of the Kobe City Eye Hospital for their ongoing support.

## Author Contributions

T. Maeda, M.Y and N.M conceived the study and performed the examinations. M.M,T. Matsuyama and M.Y analyzed and interpreted the examination data. T. Matsuyama conducted statistically analysis. M.Y, T. Matsuyama, and M.M. wrote the manuscript, which was approved by all authors prior to submission. A.M,Y.H,S.T, N.M and D.S, provided helpful guidance and suggestions. Y.K and M.M directed all work.

## Abbreviations

CF: Counting fingers
FST: Full-field stimulus testing
HM: Hand motion
IOP: Intraocular pressure
IRD: Inherited retinal disease
LP: Light perception
iPSC: induced pluripotent cell
NLP: Non light perception
OCT: Optical coherence tomography
RP: Retinitis pigmentosa
RPE: Retinal pigment epithelium

## Notes

Financial Support: Japan Agency for Medical Research and Development (AMED) under grant number JP22bm0204002

Conflict of Interest: No conflicting relationship exists for any author

### Competing Interest Statement

The authors have declared no competing interest.

### Funding Statement

This study was funded by Japan Agency for Medical Research and Development (AMED) under grant number JP22bm0204002

### Author Declarations

The study protocol was approved by the institutional review board of Kobe City Medical Center General Hospital

### Summary of Updates

Correction of title and a few minor corrections to the main text

